# Unraveling the Impact of Therapeutic Drug Monitoring via Machine Learning

**DOI:** 10.1101/2023.10.23.23297424

**Authors:** H. Ceren Ates, Abdallah Alshanawani, Stefan Hagel, Menino O Cotta, Jason A Roberts, Can Dincer, Cihan Ates

## Abstract

Clinical studies investigating the benefits of beta-lactam therapeutic drug monitoring (TDM) among critically ill patients have been hindered by small patient group, variability between studies, patient heterogeneity and inadequate use of TDM. Accordingly, definitive conclusions regarding the efficacy of TDM have remained elusive. To address these challenges, we propose an innovative approach that leverages data-driven methods to unveil the concealed connections between therapy effectiveness and patient data. Our findings reveal that machine learning algorithms can identify informative features that distinguish between healthy and sick states. These hold promise as potential markers for disease classification and severity stratification, as well as offering a continuous and data-driven “multidimensional” SOFA score. The implementation of TDM-guided therapy was associated with improved recovery rates particularly during the critical 72 hours after sepsis onset. Providing the first-ever quantification of the impact of TDM, our approach has the potential to revolutionize the way TDM applied in critical care.

---

Sepsis is a life-threatening condition that poses significant challenges to healthcare professionals due to its difficulty in early detection and management, leading to a high mortality rate. Intravenous antibiotic therapy, including the commonly used beta-lactam class of antibiotics, is a crucial element in the management of sepsis^1^. Antibiotic administration should start as soon as possible, ideally within the first hour of diagnosis and after clinical cultures are obtained^2^. Early recognition and optimized treatment of sepsis can improve the chances of patient survival. Due to the heterogeneous presentation of sepsis, however, early recognition is often challenging. This can lead to delayed care, increasing the risk of organ failure and negatively impacting patient outcomes.

Due to acute disease processes and treatment interventions associated with sepsis and its management in the intensive care unit (ICU), critically ill patients often experience altered pharmacokinetics (PK)^3–5^. This can result in highly variable and unpredictable exposures of beta-lactam antibiotics^6^. Moreover, due to antibiotic usage being higher in the ICU compared to other areas of the hospital and in the community, pathogens isolated in ICU patients are at risk of reduced antibiotic susceptibility. This adds to the difficulty in ensuring beta-lactam antibiotic exposures attain desired pharmacodynamic (PD) targets^7^. Therapeutic drug monitoring (TDM) offers a potential solution to ensure antibiotic concentrations are maintained at target exposures throughout the treatment period. This intervention may help improve treatment failure rates and reduce the risk of exposure-related drug toxicity.

However, the wider adoption of beta-lactam TDM in ICUs is impeded by several challenges, including limited availability, operational complexities that can delay turnaround times for reporting results, as well as cost considerations^8^. As a result, healthcare professionals are compelled to carefully assess the optimal allocation of resources and prioritize patient groups that are likely to derive the greatest benefits from beta-lactam TDM^9^.

Despite several randomized controlled trials investigating the impact of beta-lactam antibiotic TDM in the ICU^2,10–13^, none have yet demonstrated a significant difference in patient outcomes. To address this gap, it is important to approach the problem from a broader perspective, considering its multi-dimensional nature. This involves examining a wider range of outcomes including clinical cure, microbiological eradication, development of antibiotic resistance, patient morbidity and mortality, as well as conducting rigorous cost-effectiveness analyses. By considering and analyzing such multifaceted information, a deeper understanding of how to optimize beta-lactam antibiotic dosing strategies in critically ill patients can be obtained^5,7,14^.

However, understanding and processing such large dimensional and heterogeneous data is not straightforward with conventional methods. In this context, machine learning (ML) emerges as a powerful tool to navigate these complexities. By harnessing ML, informative features can be identified from the collected multi-dimensional, temporal patient data, enabling the creation of a comprehensive patient state representation. This learned representation facilitates the distinction between healthier and relatively sicker states by comparing informative measured features over time within a patient and across different patients. Such data-driven analyses offer the means to monitor patients’ recovery trajectories and treatment responses, shedding light on the intricate interplay between therapy, patient dynamics, and outcomes. Inclusion of ML-based methodologies thus provides a crucial lens through which to quantify the impact of TDM on patient recovery, enhancing our ability to derive meaningful insights from the intricate web of clinical data. The objective of this work is to quantitatively analyze the influence of TDM on recovery trajectories, with specific attention to three key aspects: (i) quantification of the patient state during Piperacillin/Tazobactam antibiotic therapy, (ii) the impact of Piperacillin/Tazobactam antibiotic TDM on patient state dynamics and (iii) the effect of Piperacillin/Tazobactam TDM on the survival of patients.

## Results

### Data Driven Assessment of TDM and Control Group Split

We first computed the mean pairwise Euclidian distances in the original feature space, which yielded no difference between the TDM and control group patients (Figure S2). Considering the small number of patients in the study (n=248), we further extended the similarity analysis in lower dimensional representation generated by two dimensionality reduction techniques, t-distributed stochastic neighbor embedding (t-SNE) and linear Principal Component Analysis (Figure 2a-b). In both cases, computed pairwise distance statistics and their distribution were the same. The analysis of the pathogen reports for the first day were also aligned with this data driven deduction. Figure 3d demonstrates the presence of distinct pathogen types in both the TDM and control group on day 1 (following randomization). Notably, the pathogen distributions exhibited similarity between the two groups. Furthermore, the distribution of piperacillin-resistant (Figure 3d) and sepsis-causing pathogens (Figure 3e) followed the same pattern. Therefore, it was deduced that patients in both the TDM and control sub-populations started treatment in similar conditions, enabling the conduction of proposed state space trajectory analysis objectively.

**Figure 1.**
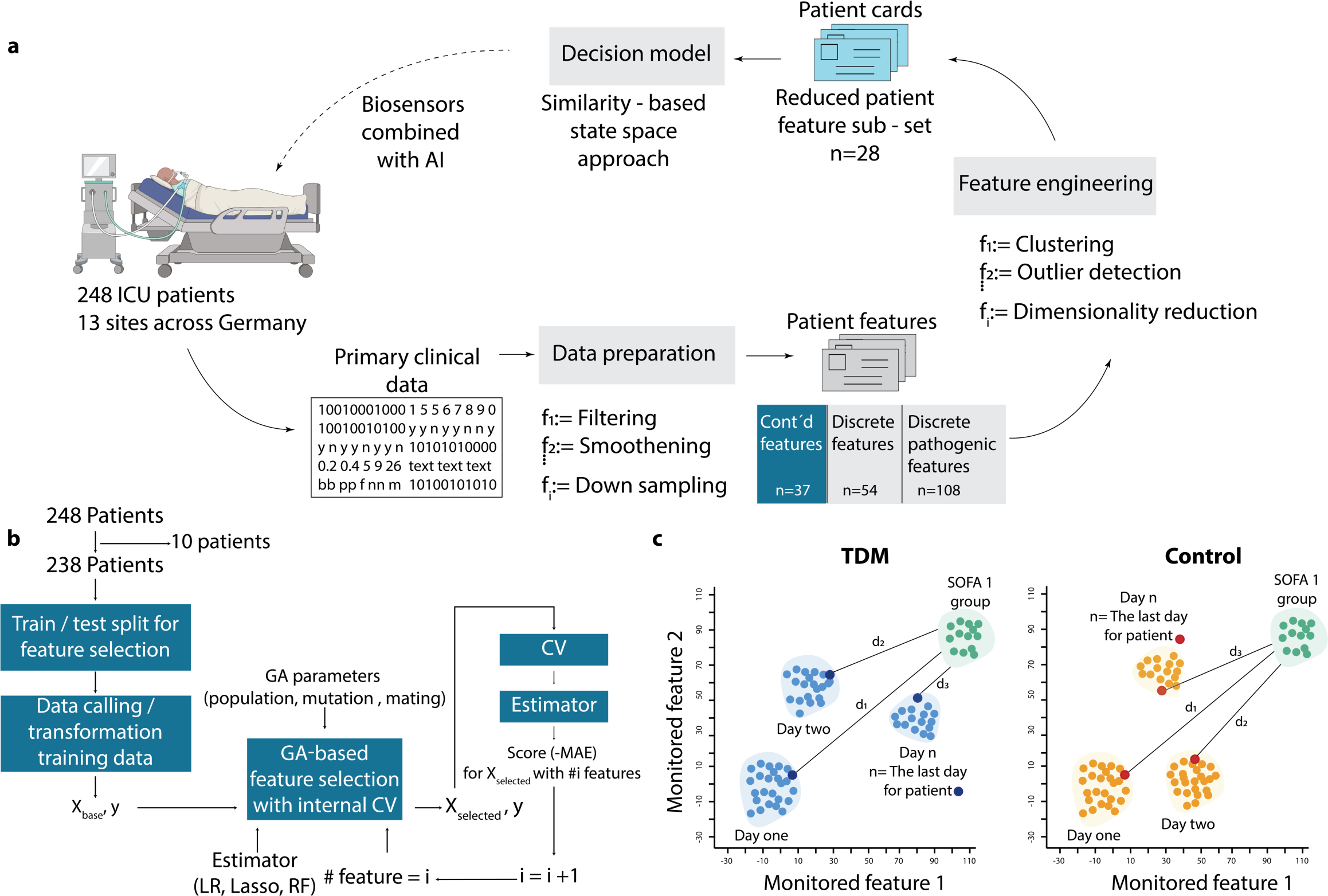
Quantifying effect of TDM by a similarity-based state-space approach. **a,** Data processing and feature selection pipeline for patient status analysis using the in-house data mining code. The heterogeneous medical database collected from hospitals is transformed into digital patient cards, followed by the selection of the top 28 representative features through feature engineering. A similarity-based state tracking approach is employed to compare the impact of TDM on patient status. The integration of biosensors for frequent sampling and enhanced drug dosage control is proposed to complete the loop and further optimize patient care. **b,** Feature selection workflow utilizing genetic algorithm (GA) implementation. The process involves leaving out 10 patients for generalizability testing, train/test split for feature selection evaluation, feature scaling/transformation, and iterative refinement with GA with cross-validation. The final feature set is determined through a frequency analysis of 100 repetitions. **c,** Visualization of travelled distance analogy in a 2D feature space to assess patient state dissimilarity. The blue and orange points represent TDM and control group patients, respectively. The distance to the reference health state (d1) indicates the degree of dissimilarity from the “healthy” state. Patients in both groups start their recovery trajectory in a specific sub-space of the 2D state space and are expected to move towards the reference state over time. The rate of recovery is determined by how quickly the groups progress from their initial states to the reference “healthy town” of Sequential Organ Failure Assessment (SOFA) score of 1. The cumulative sum of Mahalanobis distances is calculated to quantify the difference between TDM and control groups’ proximity to the healthy zone for each day.

**Figure 2.**
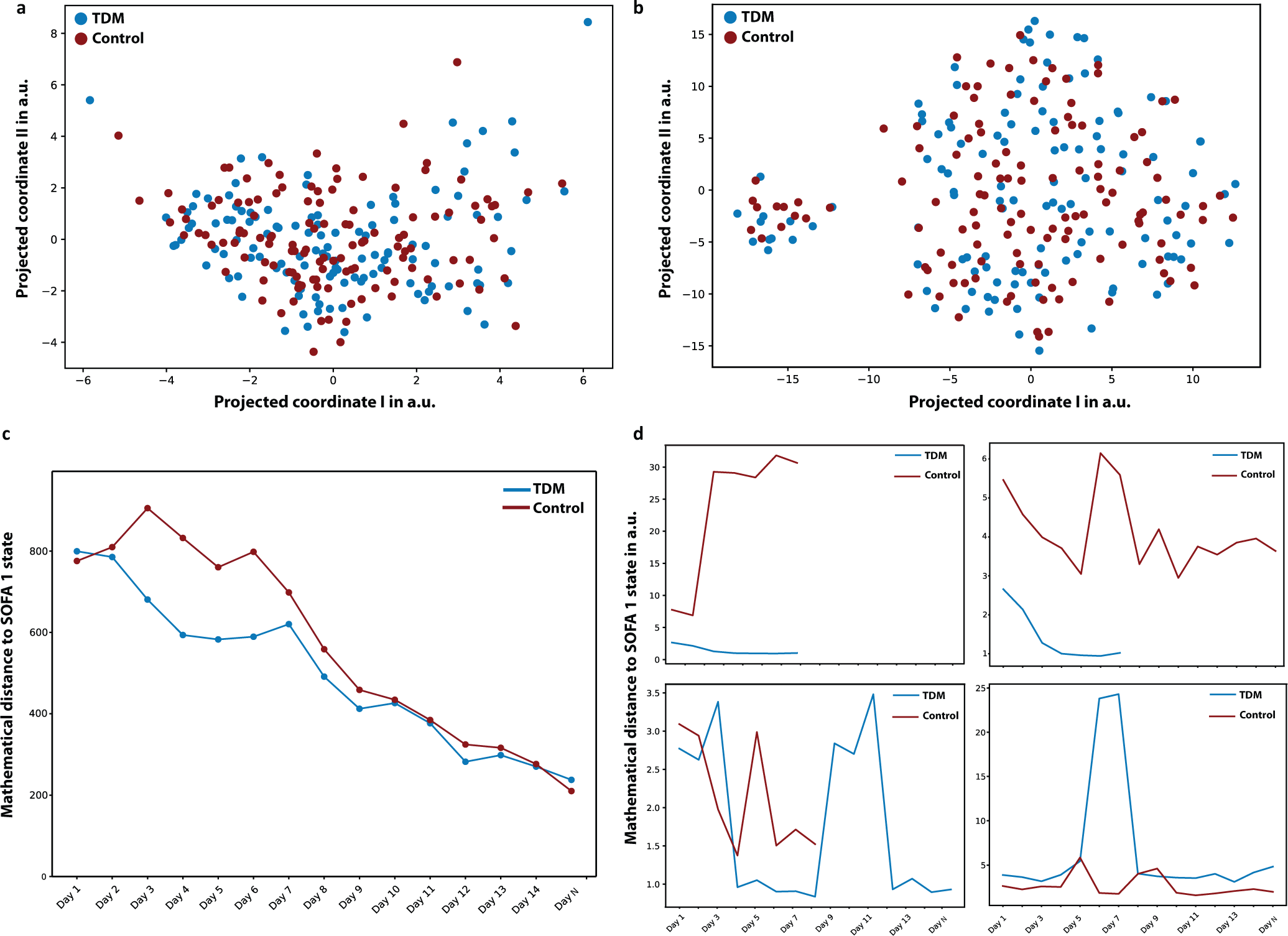
Data driven assessment of TDM and impact of TDM on patient state trajectory. The underlying hypothesis of this study is that the medical data collected daily during the clinical study holds valuable information regarding the patients’ health states. By employing mathematical techniques of similarity, the gradual changes in patient states based on the distribution of their health states were quantified. Mathematical similarity of patient groups on day 1 is demonstrated using two dimensionality reduction techniques: **a,** t-distributed stochastic neighbor embedding (t-SNE) and **b,** Principal component analysis. Both t-SNE and PCA embeddings indicate that patient states were homogenously distributed on the day of admission, validating the random TDM and control split. Quantitative comparison in high dimensional feature space is given in the Supplementary Information. **c,** The concept of “traveling to a healthier state,” which is evaluated by calculating the normalized Mahalanobis distance between the patient’s health status on each day and the reference state. **d,** Randomly sampled individual patient trajectories from both the control and TDM groups, revealing the disparity in mathematical distance towards the SOFA 1 state.

**Figure 3.**
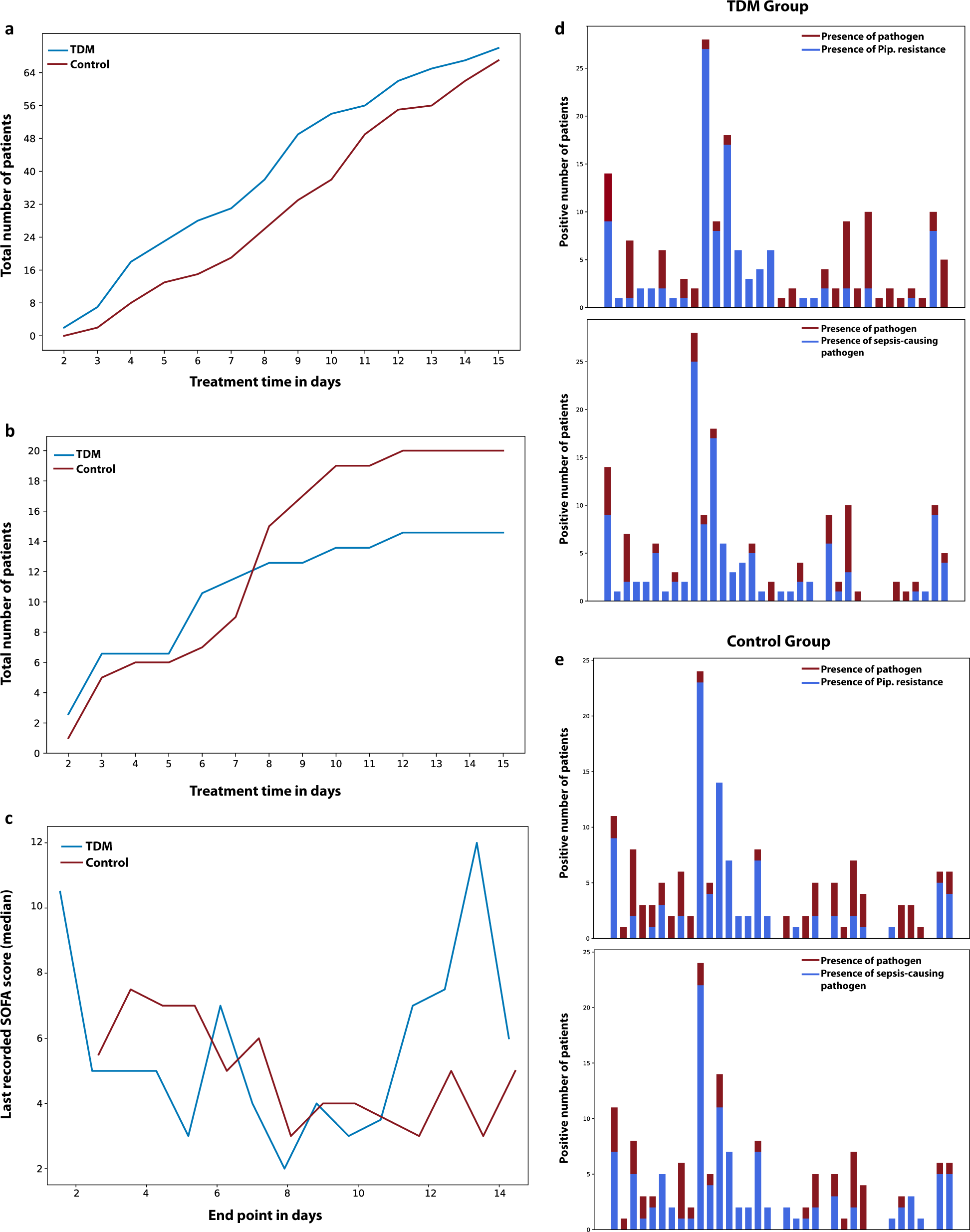
Effect of TDM on patient recovery trajectories. **a,** Number of people left the study alive in both control and TDM groups. **b,** Number of people left the study dead in both control and TDM groups. **c,** Last recorded SOFA scores (median) for patients left the study alive in both TDM and control groups. **d,** The presence of distinct pathogen types (red) with the distribution of piperacillin resistant pathogen (blue) in both the TDM and control group on Day 1 (following randomization). **e**, The presence of distinct pathogen types (red) with the distribution of sepsis-causing pathogen (blue) in both the TDM and control group on Day 1 (following randomization). Pathogen distributions, the distribution of piperacillin resistant and sepsis-causing pathogens exhibit similarities between the two groups, indicating that the patients in TDM and control sub-populations started the treatment in similar conditions.

### Impact of TDM on Patient State Trajectory

The evolution of the patient states for the TDM and control group populations revealed three important outcomes (Figure 2c). At the beginning of the study, both the TDM and control groups demonstrated a comparable proximity to the reference state space (i.e., patients with SOFA score = 1), indicating similar feature distributions within this patient group. This finding aligned with the results illustrated in Figure 2a-b, emphasizing the resemblance in patient statuses between the control and TDM groups during the initial phase of the study. Secondly, as treatment continued, the TDM groups “moved” faster towards the reference state compared to the control group. In particular, the distance between the TDM and control groups were found to be the greatest 48 and 72 hours after the randomization. It should be noted that all patient received the same dosage at the beginning of the clinical study (Day 1), and dose adjustments were made once the data is available for TDM group on Day 2. The movement after dose adjustment (Day 3 and 4) towards the reference state quantitatively demonstrated that the effective distance travelled per day was much higher for the TDM group. Thirdly, the movement of TDM and control groups to a healthier state were similar after Day 10, corresponding to the day that TDM guided dose adjustment ceased for all TDM patients. In other words, the treatment of the patients in the TDM and control groups were conducted in the same way after Day 10, and the patient state trajectories also reflected this applied procedure. These observations indicate the capability of the state trajectory approach to assess different stages of the treatment process. Moreover, randomly sampled individual patient trajectories from the TDM and control group (Figure 2d) revealed unique responses to the therapy, highlighting the individualistic nature of the treatment process and the need for an individualized therapy management for a better recovery.

### Patient Recovery Analysis

Although TDM and control patients started the therapy at similar conditions as a group, the individual states of the patients demonstrated a variance. Therefore, instead of looking at the statistics of the whole patients independent of the time frame, we examined the data of the last day recorded. The total number of patients left the study as alive (Figure 3a) was typically higher with TDM, particularly in the period of dose adjustment (between Day 2 and 10). More importantly, the slope for the curve of patients died during this study (Figure 3b) was noticeably smaller for patients with TDM-guided therapy during the dose adjustment period, resulting in a lower mortality rate. When the status of the patients leaving the study alive was further examined (Figure 3c), it is seen that the last recorded SOFA scores were also lower for the TDM group for the first 10 days, with the exception of Day 1 in which 2 patients left the study with a high SOFA score. This is also consistent with the fact that in the original statistical analysis of the study, a lower mortality rate and a higher clinical and microbiological cure rate were observed in patients receiving TDM-guided therapy, although these differences were not statistically significant.

We postulate that the observed ‘spike’ in SOFA scores between Day 12 to 14 among patients who left the clinical study alive within the TDM group can be attributed to two interrelated factors. Firstly, the remaining smaller population size in TDM group could potentially magnify the impact of individual variations in patient responses. Secondly, a relatively higher proportion of critically ill individuals remained in the TDM group compared to the control as a result of the TDM group’s overall lower mortality rate throughout the first 10 days.

## Discussion

TDM is the practice of measuring, analyzing and adjusting the drug levels in a patient’s blood to achieve the desired therapeutic outcomes while avoiding adverse effects. Herein, the drug concentrations measured at specific intervals provide valuable information about a patient’s individual PK/PD, allowing the healthcare providers to tune treatment regimens based on the individual state and make “informed” decisions regarding dosage adjustments or changes in medication. Therefore, the success of the TDM practices strongly depends on how frequently and accurately the drug concentrations are measured, and how the patient current state is correlated with the therapeutic window. Our study utilizes data collected in a clinical trial conducted across 13 sites, where half of the patients were subjected to dose adjustment. Notably, our findings demonstrate the capability of machine learning algorithms to discern meaningful features, allowing us to quantitatively capture the patient recovery process and gain valuable insights into therapy progression.

Despite the rich clinical data regarding patient state (199 features), however, the dynamic TDM process was only run using a daily granularity. That is, the dose adjustment in the beta-lactam TDM group was made once a day, thus smaller time interval fluctuations in drug concentrations could not be captured. Likewise, of the piperacillin concentrations measured in the TDM group, 88.1% (n = 510) were reported on the same day, while 10.0% (n = 58) were reported at a later date and 1.9% (n = 11) were never reported^2^. As a result of this “process delay”, TDM feedback cycle used to adjust the dosage regimen was potentially slower when compared to the evolution of the patient state, which can be seen in the daily SOFA score fluctuations (Figure S6). Another limitation of the clinical study was the limited number of dosage adjustments in the TDM group. Hence, the dose adjustment may not have been made at the optimum time for the patients. For instance, at day 1, for only 70% of the TDM patients dose adjustment was considered necessary, followed by an average of 48% during treatment, and ended with less than 30% for Day 9 and Day 10. Four patients in TDM group had never received a dose adjustment^2^.

The other limitation of the TDM workflow applied was its dependency on the minimum inhibitory concentration (MIC)^2^, which represents the lowest concentration of a drug that is required to inhibit the growth of the microorganism in a laboratory setting. In the TDM patients, the dose adjustment was based on the most recent pathology report and the corresponding MIC values. Nonetheless, setting the therapeutic window based on MIC has two important limitations: (i) serum drug concentrations do not necessarily represent the concentration at the site of infection, hence the drug concentrations above MIC may not been reached at the site^15^, (ii) complex PK/PD of the patient status may demand a drug concentration different than the MIC. As MIC value only distinguishes between growth and suppression of pathogen under the lab conditions, the drug concentration recommended by MIC may not be sufficient to kill the pathogen at the site of infection. In an ideal scheme, MIC values can be used to set the initial therapeutic window, which is continuously updated based on an individualized patient model stemming from the site of infection^8,16^. However, despite these challenges, ML-augmented analysis of the patient data conclusively demonstrated that implementing dose adjustment policies had a significant and favorable impact on the overall recovery of patients in the TDM group. In particular, dose adjustment based on TDM had the largest impact within the first 72 hours of admission, which is noted as an important treatment window in the management of patients with sepsis.

By quantifying the effects of TDM, it becomes evident that aligning antibiotic exposure with the patient’s instantaneous state has the potential to optimize treatment outcomes. In the clinical case being examined, for instance, updating patient data on an hourly basis, rather than daily, could significantly amplify the impact of TDM, especially within the first 72 hours. In this regard, use of biosensors can potentially provide accurate, on-site and rapid detection of the drug concentration from both blood and non-invasive bodily fluids including sweat, saliva, tear and breath, providing a chance to accelerate the response time of the TDM cycle^17–22^. Additionally, this sensor data can be utilized to build digital patient models, which can further enable model predictive control policies via simulating the patient response with individualized PK/PD parameters. Herein, trainable model parameters can be learnt during therapy with the rapid, continuous stream of sensor-based measurements of drug concentrations. By integrating population-specific antibiotic PK models with patient-specific information such as kidney function, weight, pathogen data, and TDM results, tailored dosing regimens can be calculated via model informed precision dosing (MIPD) software ^8,16,18^. Preliminary findings indicate that such a personalized approach improves the attainment of PK/PD targets, particularly for patients at high risk of mortality from infections^23^.

In therapeutic drug management, it is vital to quantify the impact of the defined drug concentration on a patient’s recovery trajectory. The analysis provided in this study can facilitate the identification of the optimal free drug concentration for a specific individual during a particular phase of treatment. By doing so, it would be possible to determine the most effective drug concentration to administer, ensuring the best possible outcomes for the patient. This, however, requires a quantitative description of the patient state. In this study, we showed that improvement in the patient state can be quantified, if the proper measured features are available. Recent developments in the wearable body area network^24^, where multiple wearables mounted on different parts of the body to concurrently analyze various physiological markers, and the integration of internet of things (IoT) devices into healthcare monitoring^25^ are enabling such continuous feature collection possible. By leveraging ML capabilities, voluminous sensor data, together with the previous medical records, genetic information and individualized healthy reference state, data driven algorithms can continuously adapt and refine dose adjustment strategies and the pharmacokinetic models in real time.

Another key role that wearable technology can play is the definition of an individualized reference healthy state. Our analysis showed that patient states can be defined from measured physiological parameters and observations, and the effect of therapy can be quantified by comparing the instantaneous patient state with a reference “healthy state”. In the current implementation, we used the SOFA=1 patient states as reference distribution, and closeness to this reference distribution provided a proxy measure for “being healthy” for all patients. In a 4P (predictive, preventive, personalized and participatory) medicine concept enabled by wearable sensors^24,26^, individualized healthy states for patients can be learned, which would further increase the accuracy of state space tracking of health status.

The proposed analysis in this study reveals an intriguing finding regarding the clinical relevance of the selected feature set and its relationship to the SOFA score. The features commonly used to calculate the SOFA score in clinical practice, such as thrombocytes, urine, creatinine, GCS, and encephalopathy, were also found to be informative in this study. From a data science perspective, the calculation of the SOFA score can be viewed as a rule-based technique that reduces the dimensionality of the data, transforming a 28-dimensional vector into a single scalar value. This interpretation leads to two practical outcomes. First, it highlights the need to understand the limitations of the SOFA score analysis and how it can be interpreted. Second, it sheds light on the proposed state space patient trajectory analysis, which can be seen as a high-dimensional, continuous version of the SOFA score.

At the first glance, reduction of all clinically relevant data as the SOFA score can be considered as a practical way for interpretation. However, the way SOFA scores are calculated in current practice can result in inaccurate clustering or classification of patients with different status. The SOFA score assesses the functionality or degree of failure in six key systems: respiratory, cardiovascular, hepatic, coagulation, renal, and neurological. It’s important to recognize that while two patients may have the same SOFA score, their clinical situations differ significantly. For instance, one patient may have impaired renal function resulting in their score of 4, whereas another patient may experience minor issues across four different major systems. Therefore, interpreting SOFA scores requires careful consideration of the specific organ systems involved to obtain a comprehensive understanding of each patient’s condition.

To address these limitations, our analysis provides a way to maintain a “multidimensional” SOFA score based on Euclidean distances. This allows for a quantitative assessment of the mathematical similarity between patient states while preserving the maximum amount of information. Hence, the method proposed in this study can be interpreted as a continuous and extended form of the SOFA score analysis. By utilizing this approach, it becomes possible to distinguish between patients with the same SOFA score but different physiological states. This enhanced level of differentiation provides valuable insights for clinical decision-making and patient management.

Our study has revealed significant findings with implications for ML-augmented disease classification, patient stratification, and monitoring treatment response. Features identified with ML techniques accurately reflected the recovery process of sepsis patients in ICUs, providing valuable insights into therapy progression and effectiveness. Importantly, continuous monitoring of these features enabled precise measurement of the recovery rate, emphasizing their potential as indicators of treatment response. By using artificial intelligence, we demonstrated for the first time quantitively that beta-lactam antibiotic TDM implementation leads to higher recovery rates and optimized patient outcomes. Our findings highlight that the state between healthy and sick individuals can be differentiated from the temporal data, which in turn can be used to quantify the recovery process as a reliable measurement of the recovery rate. Additionally, TDM-guided dosing was found to significantly alter the trajectory of recovery, underlining its potential for personalized medicine and enhanced patient care.

## Data Availability

All data produced in the present study are available upon reasonable request to the authors.

## Acknowledgements

H.C.A and C.D. would like to acknowledge the Deutsche Forschungsgemeinschaft (DFG, German Research Foundation) for funding this work (grant numbers 404478562 and 446617142). The TARGET study was funded by the Federal Ministry of Education and Research (BMBF), 01EO1502. TARGET Trial Investigators: Friedhelm Bach, Thorsten Brenner, Hendrik Bracht, Alexander Brinkmann, Thorsten Annecke, Andreas Hohn, Markus Weigand, Guido Michels, Stefan Kluge, Axel Nierhaus, Dominik Jarczak, Christina König, Dirk Weismann, Otto Frey, Dominic Witzke, Carsten Müller, Michael Bauer, Michael Kiehntopf, Sophie Neugebauer, Thomas Lehmann, Jason A. Roberts, Mathias W. Pletz on behalf of the TARGET Trial Investigators.

## Methods

The data source used in this work involved patients admitted with severe sepsis or septic shock and aimed to compare the clinical effectiveness of TDM-guided Piperacillin/Tazobactam antibiotic therapy versus a fixed dosing strategy. The working hypothesis here is that the information regarding the recovery process is embedded into the measured features. In other words, healthier states should be distinguishable from relatively sick states by comparing informative measured features at different times for a patient, or in between patients. Such an approach converts the problem of quantifying the effect of TDM into a state trajectory analysis; that is, via monitoring the change in these features, recovery rate can be measured in the state space. More importantly, it becomes possible to quantify relative recovery rates with or without TDM, as a more effective therapy will change the recovery trajectory for the patient.

### Details of the clinical study

A clinical trial was conducted to compare the effectiveness of TDM-guided antibiotic therapy with fixed dosing in improving clinical outcomes in sepsis patients treated with piperacillin/tazobactam^2^. The trial included 248 adult patients with severe sepsis or septic shock who had received the therapy within the last 24 hours before enrollment. It took place in 13 different locations in Germany between January 2017 and December 2019 and was randomized, controlled, and patient blinded.

The study recorded clinical, microbiological, and laboratory data from the day prior to randomization and then throughout the following time points: day 14 post randomization, at the end of therapy, at discharge from the ICU, and at day 28. Patients were randomly assigned (1:1) to either the TDM group or to the control group (no-TDM). Following randomization, both the control and TDM groups were given an initial loading dose of 4.5 g of piperacillin/tazobactam, followed by a continuous infusion of the same antibiotic. The total daily dose was 13.5 g (9 g in patients with an estimated glomerular filtration rate (eGFR) < 20 mL min^-1^). In the TDM group, dosing of piperacillin/tazobactam was guided by daily monitoring of piperacillin, starting on Day 1 post randomization (or Day 0 if the piperacillin concentration had already reached a steady state) for a maximum of 10 days. Use of antimicrobial combination therapy, termination or (de-) escalation of antimicrobial therapy was allowed at any time and at the discretion of the treating physicians. The target plasma concentration of free piperacillin was set to four times (with a range of ±20%) the minimal inhibitory concentration of the pathogen responsible for sepsis. In patients receiving TDM-guided therapy with piperacillin/tazobactam, a dose adjustment was made on 53.9% (312/579) of the treatment days. In the control group, daily dose adjustments were based on patient renal function and did not utilize any TDM. Both patient cohorts had blood samples taken daily to measure piperacillin concentrations. The TDM group received same-day analysis, reporting, and dose adjustments, while analysis in the control group could be performed on the same day or later, with samples kept at - 80°C until analyzed. Total piperacillin concentration measurements were performed on-site in study centers using either high-performance liquid chromatography (HPLC) or liquid chromatography mass spectrometry (LC–MS/MS). The trial protocol was approved by institutional review boards, published previously, and Germany’s Federal Institute for Drugs and Medical Devices (EudraCT: 2016-000136-17, ref: 4041358). The primary endpoint was sepsis-related organ dysfunction measured by the mean daily total SOFA scores over 10 days, discharge from the ICU or death, whichever occurred first. The mean SOFA score was calculated as the mean of all daily SOFA scores for each patient.

In the clinical trial no significant beneficial effect of TDM-guided piperacillin/tazobactam therapy with regard to the 10-day mean total SOFA score compared to fixed dosing was observed (7.9 points (95% CI 7.1–8.7) in the TDM group vs. 8.2 points (95% CI 7.5–9) in the control group, in between group difference 0.3 points, 95% CI − 0.4 to 1, *p*=0.39). However, patients who underwent TDM-guided therapy displayed a 4.2% lower 28-day mortality and a higher rate of microbiological and clinical cure during therapy, but these differences were not statistically significant.

### Patient state and measure of similarity

Mahalanobis distance is a statistical measure used to assess the dissimilarity between a sample point and a distribution in a multidimensional space, considering the structure of the data. The Mahalanobis distance from a patient state vector A to a reference distribution R with mean μ and covariance Σ is calculated as:

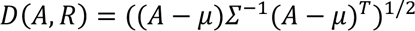

Where Σ^1^’ is the inverse of the covariance matrix of the reference distribution R. The Mahalanobis distance accounts for the correlation between different measured variables by scaling the differences with the inverse covariance matrix^27^. Considering the inter-patient variance in the measured physiologically relevant features, it is considered that the Mahalanobis distance would be the best fit to describe the dissimilarities between the patient states.

In particular, we used the Mahalanobis distance to measure dissimilarity between the patient state at a given time and a reference “healthy state distribution” based on the SOFA score. Firstly, we investigated the uniqueness of the SOFA scores during the Exploratory Data Analysis (EDA) phase, revealing that only six patient states have a score of SOFA = 0. As a result, second best SOFA score, SOFA = 1 is used as a filter to create a state vector group as the reference distribution. Figure 1c depicts the measurement of Mahalanobis distances for each patient state at each day in a 2D feature space. Herein, blue and orange points mark the state vectors of the patients in a 2D state space for the TDM and control groups, respectively. In both groups, patients start their recovery trajectory at a certain sub-space of the 2D state space. The distance to the reference health state (for example, d_1_) is expected to be correlated with the degree of dissimilarity with the state of “being healthy”. As the therapy continues successfully, the patient should “move” in the feature space towards the reference state and the rate of recovery is correlated with how fast the groups move from their initial states (day of admission) to the reference zone, so called “healthy town”. In other words, if the dose adjustment within TDM is beneficial for the TDM group, there must be a distinct difference between how much closer they are to the healthy zone compared to the control group. This is quantified by calculating the cumulative sum of the Mahalanobis distance between the TDM/control groups and the reference states for each day (t):

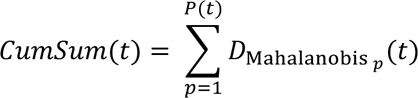

Where p is the patient for which the distance (*D_Mahalanbisp_*) is being calculated for day t and *P(t)* the number of patients that are in the TDM/control group at day t. To ensure the statistical significance of the measured CumSum, daily values are normalized based on the mean pairwise distance of SOFA = 1. In other words, normalized distances shown in Figure 3c report how far away the TDM/control group to the reference state, if the distance between SOFA = 1 patients is equal to 1.

It should be also noted here that since the feature set is heterogenous (i.e., consists of continuous, categorical and ordinal variables), discrete ones should be first transformed into a pseudo-continuous representation, and then all features should be scaled for an unbiased dissimilarity analysis. In this work, the features are first grouped into three sub-sets: continuous, discrete and discrete pathogen-related features (see Supplementary Information). Then, discrete features are transformed via CatBoost^28^ to have a continuous feature space. Next, all features are normalized with standard scaling before conducting the distance analysis. CatBoost and standard scaling methods are fit by using only the training data to prevent data leakage.

### Feature engineering and selection

Mathematical similarity, which is type of unsupervised learning approach, relies on the measured distances in high dimensional feature space, which makes feature engineering and selection very critical, particularly for the conducted analysis with limited number of patients and total number of daily observations (state vectors)^28^. In the current study, the primary clinical data is first analyzed in terms of feature variance, missing data, outliers, and any other unphysical abnormalities (Supplementary information). The data preparation steps (Supplementary Information) converted primary clinical data into 199 structured features for 248 patients (TDM:123, control:125) with variable trajectory lengths (i.e., duration of the treatment at daily granularity), consisting of 2376 state vectors in total. Since the clinical data is limited, high dimensional feature space (199 dimensions) is found to be extremely sparse, hence it has to be reduced to alleviate the dimensionality problem. In particular, the dimensionality of the problem has to be reduced, as the similarity analysis relies on distance, and the “signal/noise ratio” in distance-based (or error-based) approximations diminishes exponentially as the number of features increases^28^. Therefore, at the next step, we applied alternative approaches including dimensionality reduction, feature selection via filtering, implicit and wrapper methods. For this particular problem, using genetic algorithms (GA) for feature selection^29^ (i.e., as a wrapper) provided the best feature subset^30^, and the methodology for feature selection is described here only for the GA implementation.

Firstly, we leave 10 patients out of the feature selection study to increase and test the generalizability of the similarity approach. The feature selection procedure is shown in Figure 1b. The process starts with a train and test split for the selected 238 patients. The training data is then passed to the Wrapper, which is a GA implementation with internal cross-validation (CV). A random forest model^31^ was used as the estimator of the feature selection wrapper. The score needed to iteratively refine the feature subset is taken as the SOFA scores, where the metric for the fitness and CV is selected as the negative mean absolute error. The number of feature subset is also scanned parametrically, starting from one to the maximum number of features to analyze the value of added information with increased number of features. The CV scores are then examined to determine the optimum number of features. As the GA involves randomness, the whole process is repeated 100 times, where 100 generations are created at each run. The frequency of the features selected by the last GA generations is given in Figure 4. It is seen that pathogen related data was rarely selected by the model, typically less than 10% of the time. For the final set, the features that were picked more than 10% of the time were unionized with a new GA iteration to cover potential multivariate correlations (Figure 4a-d, dark blue). More details about the feature selection scores are presented in Supplementary Information.

**Figure 4.**
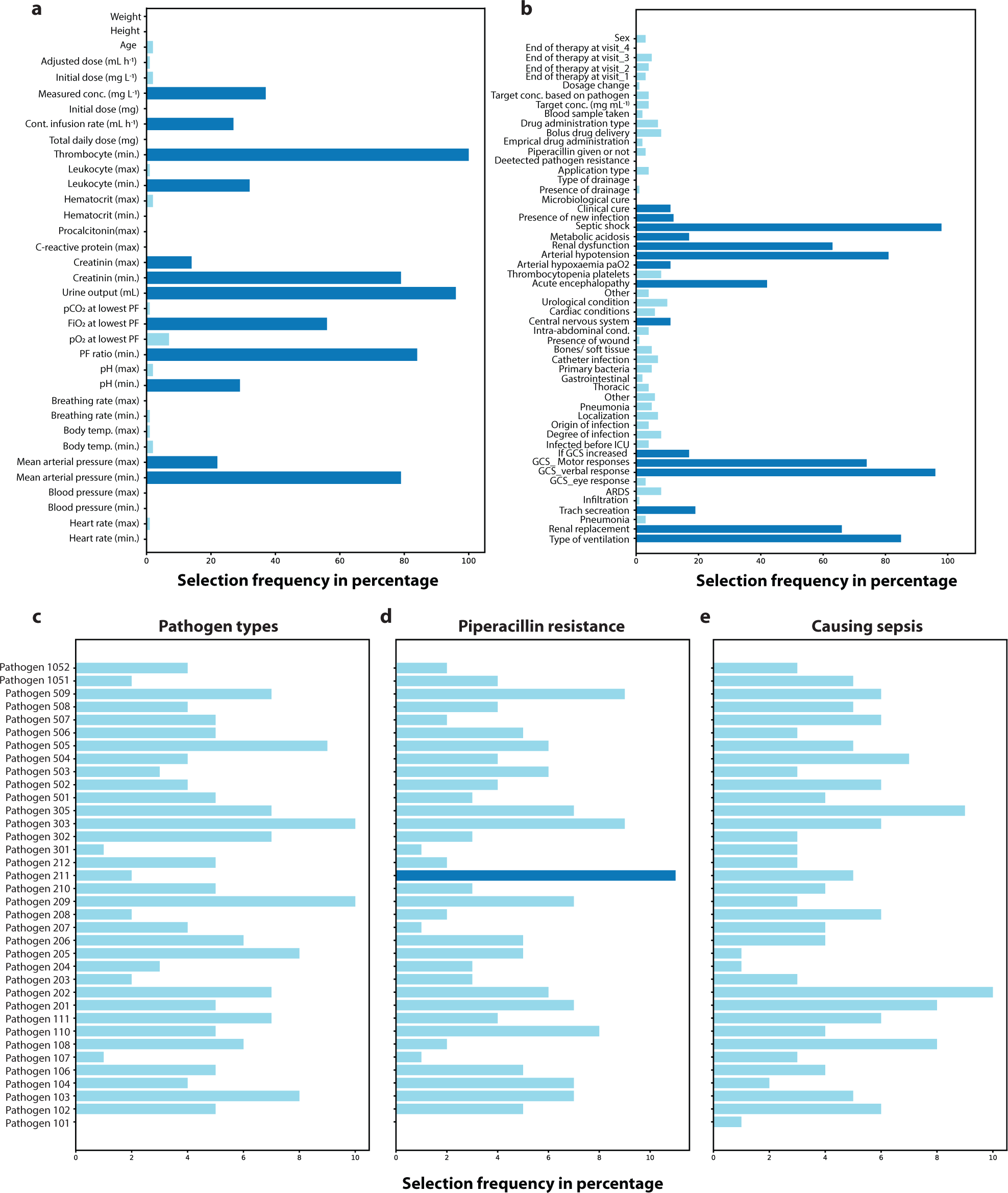
Features considered relevant by the evolutionary feature selection algorithm. Each bar denotes how many times a feature was picked by GA for SOFA score prediction. Features used in the patient state analysis is highlighted as dark blue. **a,** Continuous features encompass various demographic information (age, height, body weight), laboratory (leukocyte count, hematocrit levels, creatinine levels), drug-related information (concentration, infusion rate), and physiological measurements (such as breathing rate, body temperature, and mean arterial pressure). **b,** Discrete features consist of yes/no questions and ordinal variables, such as the presence of metabolic acidosis, renal dysfunction, or the need for renal replacement. **c,** Whether, a pathogen could be detected in the patient. Microbiology reports cover 36 different pathogens, including gram-positive and gram-negative bacteria, fungi, and other pathogens such as Chlamydia species; **d,** Whether, a detected pathogen is resistant to piperacillin and **e,** Whether the pathogen type is responsible for the sepsis episode.

### SOFA score and patient analysis

The cumulative Mahalanobis distance analysis described in the previous section provides a way to quantify the impact of TDM on the patient status in the form of mathematical dissimilarity between the current state and a reference distribution. To further analyze how the abstract distances translate into patient recovery, we examined the (i) temporal evolution of the patient SOFA scores, and (ii) mortality rate for both TDM and control groups.

The SOFA score is a clinical tool utilized to evaluate the severity of illness and prognosis in critically ill patients. It involves assessing six organ systems: respiratory, cardiovascular, hepatic, coagulation, renal, and neurological. Each organ system is assigned a score ranging from 0 to 4, where higher scores indicate more severe dysfunction. The individual scores for each organ system are summed to obtain a total SOFA score, which can range from 0 to 24. A higher score indicates more pronounced organ dysfunction and a poorer prognosis for the patient. In the current study, SOFA scores are used in the supervised ML methods for feature selection, and to interpret and discuss the calculated Mahalanobis distances. We also conducted statistical analysis on the distribution of patient health status by examining the multivariate feature distributions on the day of admission, as well as on the last recorded patient data (state vectors). Patient analysis includes the comparison of the first day state vectors of the patients to justify the controlled clinical trial with dimensionality reduction techniques qualitatively and the Mahalanobis distances quantitatively. Pathogen tests of the TDM and control groups are also compared for the first day to ensure that TDM and control split of the patients is not biased towards any group. In other words, the pathogen distributions and their piperacillin resistances should also be distributed in a balanced way between the TDM and control group patients. Furthermore, for each day of the therapy, medians of the last recorded SOFA score for alive patients and the day patients leave the clinical study are extracted from the log files to discuss the results obtained by the similarity-based state analysis.

## Code availability

The code utilized in this study was tailored to the data collected in the clinical study and its unique data structure. As the code has little use without access to the data and, as such, has not been made publicly available. All data processing and modelling were conducted on Python 3 using standard libraries that are publicly available: *pandas, numpy, scipy, scikit-learn, matplotlib, seaborn, plotly, category-encoders, deap, sklearn-genetic, statsmodels*.

