## Supporting Information for "Unraveling the Impact of Therapeutic Drug Monitoring via Machine Learning"

### Data Preparation Procedure

There were 12 files constituting the primary clinical data. In each, the patient data is encoded as follows:

**rno: Ta-01-0001-3**

- 01: which site/hospital
- 0001: patient number

enabling to filter and process patient specific features, and create virtual patient cards. The content of each data is summarized below:

**dm:** Demographic information. It includes features like age, sex, height, body weight, hospital admission date, ICU admission date, type of assignment, where before ICU.

**vs:** Physiological data such as PF Ratio (paO2 / FiO2 ratio; only arterial measurement, lowest paO2 +associated FiO2)).

**pip:** Piperacillin therapy related data.

**lb:** Lab measurements are reported in this document.

**mibi:** Microbiology lab results.

**sep:** Sepsis related file. It contains information about site of infection, if sepsis happened and response to drug.

**amic:** Antimicrobial therapy related data.

**amyk:** Antifungal treatment related data.

**rand:** Tells which patient gets the TDM and which one is the control group.

**chir:** Surgical drainage data.

**ccmc:** Clinical cure data. Most important column is the clinical cure. The rest is considered as supplementary data.

**com:** Additional comments for some patients. Not used in this work.

Data preparation pipeline starts with data cleaning. All primary files are treated individually to exclude uninformative, redundant features. Furthermore, if there is more than one entry for a feature at a given day, they are filtered and saved as a list at the first step. In other words, each patient has one entry for each feature, at every day where the length of the entry may vary from patient to patient, feature to feature, or day to day. In the first iteration, all information filtered is kept as lists of entries for each day. Below, the decision made during the data cleaning is summarized:

- For any feature, in any file, if the values are saved with different units, they are all converted into the same unit (e.g., if majority is reported as mg, all reported as mg and g information is converted to mg).
- A new “dead” feature is created. The entry implying that patient is dead or in palliation (['Tod', 'Patient verstorben', 'verstorben', 'Versterben', 'Pat. verstorben', 'Tod des Patienten', 'Palliation, Exitus', 'Palliation', 'Death', 'Todesfall']) was not consistent and thus, we used filters to assign numerical values of 0 and 1 for alive and dead patients.
- underscoreX: Some features (e.g., START_X) denote whether corresponding feature has a missing value or not. As the information is already in the feature itself, underscore columns are dropped.
- ICU day: Since the patients are taken in ICU shortly after hospital admission, the feature had little variance in between patients, and dropped accordingly.
- Zero variance features are dropped, such as “zksno”.
- We drop features with no information, or those repeat the information given in the other features: where_before_ICU, site, time2, dsno, KONZENT_X new_dosage_day, VISITE_D_new, START_D_new,START_Z_new, STOPP_D_new, STOPP_Z_new, KONZENT_Z_new, STOPP_Z_new, LARATENEU_D_new, LARATENEU_Z_new UNIT_C (after unit conversions), WISTOFF_E, WISTOFF_C, ANTIMYK_B, hearthrate_NA, PF_unit(1:kPa-2:mmHg), new_dosage_day, comment_why_pip_changed, pip_mic_na, other_pathogen, 'Randomization_day', 'Randomization_time', 'RANDOMNR_C'
- We also remove rows with ANTIMYK_B==1, as it contained no information.
- From the clinical cure data, only the columns 'Clinical_cure(1:healing-2:improvement-3:failure-9:na)', 'Microbio_cure(1=>7-9)' are kept.

At the next step, the clean 11 data files that are saved in a different directory is read and combined under a single pandas dataframe . Herein, each row represents a patient, and each column contains variable length lists of features. The shape of the dataframe at this step was (254,152), denoting 152 features for 254 patients. First 11 column of the data is depicted below to demonstrate the df structure.

| rno | Visite_day | heathrate_min | heathrate_max | blood_pressure_min | blood_pressure_max | mean_art.bloodpressure_min | mean_art.bloodpressure_max | temp_min | ... | Height | Weight |
|---|---|---|---|---|---|---|---|---|---|---|---|
| Ta-01-0001-3 | ['2017-01-26', '2017-01-27', '2017-01-28', '20... | ['[46.0, 59.0]', '[56.0]', '[56.0]', '[46.0]',... | ['[116.0, 113.0]', '[110.0]', '[86.0]', '[94.0... | ['[63.0, nan]', nan, nan, nan, nan, nan, nan, ... | ['[121.0, nan]', nan, nan, nan, nan, nan, nan,... | ['[46.0, 64.0]', '[64.0]', '[69.0]', '[59.0]',... | ['[86.0, 83.0]', '[81.0]', '[86.0]', '[81.0]',... | ['[32.2, 34.6]', '[35.4]', '[35.7]', '[36.5]',... | ... | 170.0 | 60.0 |
| Ta-01-0002-1 | ['2017-01-27', '2017-01-28', '2017-01-29', '20... | ['[76.0, 80.0]', '[83.0]', '[72.0]', '[71.0]',... | ['[115.0, 96.0]', '[129.0]', '[105.0]', '[95.0... | ['[81.0, nan]', nan, nan, nan, nan, nan, nan] | ['[122.0, nan]', nan, nan, nan, nan, nan, nan] | ['[56.0, 65.0]', '[59.0]', '[58.0]', '[58.0]',... | ['[77.0, 79.0]', '[82.0]', '[87.0]', '[83.0]',... | ['[34.3, 37.1]', '[36.8]', '[36.9]', '[37.2]',... | ... | 164.0 | 60.0 |
| Ta-01-0003-9 | ['2017-03-22', '2017-03-23', '2017-03-24', '20... | ['[88.0, 94.0]', '[79.0]', '[76.0]', '[67.0]',... | ['[174.0, 129.0]', '[112.0]', '[98.0]', '[80.0... | ['[86.0, nan]', nan, nan, nan, nan, nan, nan, ... | ['[149.0, nan]', nan, nan, nan, nan, nan, nan,... | ['[62.0, 62.0]', '[60.0]', '[73.0]', '[79.0]',... | ['[91.0, 112.0]', '[92.0]', '[107.0]', '[111.0... | ['[36.9, 37.0]', '[36.8]', '[37.1]', '[37.2]',... | ... | 170.0 | 140.0 |
| Ta-01-0004-6 | ['2017-03-24', '2017-03-25', '2017-03-26', '20... | ['[67.0, 76.0]', '[58.0]', '[65.0]', '[81.0, n... | ['[142.0, 118.0]', '[109.0]', '[105.0]', '[133... | ['[79.0, nan]', nan, nan, nan, nan, nan, nan, ... | ['[192.0, nan]', nan, nan, nan, nan, nan, nan,... | ['[61.0, 61.0]', '[53.0]', '[59.0]', '[67.0, n... | ['[133.0, 97.0]', '[102.0]', '[96.0]', '[114.0... | ['[36.4, 37.1]', '[36.2]', '[37.7]', '[37.1, n... | ... | 185.0 | 80.0 |
| Ta-01-0005-4 | ['2017-04-20', '2017-04-21', '2017-04-22', '20... | ['[88.0, 84.0]', '[62.0]', '[60.0]', '[60.0]',... | ['[178.0, 110.0]', '[87.0]', '[73.0]', '[89.0]... | ['[78.0, nan]', nan, nan, nan, nan, nan, nan, ... | ['[129.0, nan]', nan, nan, nan, nan, nan, nan,... | ['[50.0, 56.0]', '[56.0]', '[56.0]', '[58.0]',... | ['[76.0, 72.0]', '[84.0]', '[83.0]', '[106.0]'... | ['[37.7, 36.5]', '[36.5]', '[36.4]', '[36.8]',... | ... | 155.0 | 55.0 |

**Figure S1.** Example of a master dataframe for patient records. Only the first 11 features of 5 patients are shown in the table.

Next, we compute the length of each cell in the df to decide the time granularity of the state space analysis. Extracting the lengths of features for each patient and for each day revealed that the study can be conducted only at daily granularity. As a result, features that are recorded multiple times in some patients (in some days) are further processed.

- Columns that contain no information are dropped.
- For missing values (less than 5% column wise), numerical features are imputed with the column median value, while categorical values are filled with the most frequent one.
- For the following features, if there is more than one measurement, daily average is used to represent the patient state:

```
'heathrate_min', 'heathrate_max', 'mean_art.bloodpressure_min', 'mean_art.bloodpressure_max',
        'temp_min', 'temp_max', 'breathing_rate_min', 'breathing_rate_max', 'ph_min', 'ph_max',
        'PF_ratio_min', 'pO2_at_lowest_PF_ratio', 'FIO2_at_lowest_PF_ratio', 'urine_output(mL)',
         'urine_collection(h)', 'kreatinin_min', 'crp_max', 'pct_max', 'leukocytes_min',
'leukocytes_max',
        'thrombocit_min', 'pip_dosage(mg)', 'kreatinin_max', 'contd_inf_rate(mL/h)',
'sample_conc(mg/L)',
                'new_dosage(mL/h)', 'bicarbonate_min', 'bicarbonate_max', 'pip_dosage(mg).1',
'target_conc(mg/L)'
```

- Features that include explanations or notes taken during the clinical study is dropped, as there were not sufficient examples for text mining:

```
'Stop_reason(why_8)', 'STARTGRUND_C', 'STOPPGRUND_C', 'reason'
```

- Columns start with the following is dropped as they contain repeated time information:

```
'rno', 'STARTZ_Z', 'STOPPZ_Z', 'STOPP_D', 'Stop_time', 'Visite_', 'day', 'hour', 'Date', 'date',
'KONZENT_D_new'
```

- Following columns are reorganized, before one hot encoding:

```
'type_of_ventilation(0:n-1:noninvasive-2:invasive)', 'renal_replacement(0:n-1:y)', 'Pneumo(0:n-1:y)',
'Trach_secration(0:little-1:abundant-2:ab_with_prulent)', 'Infiltrate(0:n-1:diffuse-2:localized)',
'ARDS(0:n-1:y)',
'eye_response(4:spontaneous-3:after_prompt-2:on_pain_sti-1:no)', 'Verbal(5:clear-4:confused-
3:single_words-2:single_sounds-1:no)',
'Motor(6=>1_getting_worse)', 'GCS(1:raised-2:estimated)', 'Drainage(0:n-1:y)', 'pip_give(0:n-1:y)',
'pip_give_type(1:empric-2:targetted)', 'Bolus_delivery(0:n-1:y)', 'blood_sampling(0:n-1:y)',
'Drainage_number', 'new_dosage_reason(1:TDM-2:side-effect-3:other)',
'Clinical_cure(1:healing-2:improvement-3:failure-9:na)', 'Microbio_cure(1=>7-9)', 'change_in_pip(0:n-
1:y)', 'why_therapy_stop(1=>8)', 'CHECK1_B(0:n-1:y)', 'CHECK2_B', 'CHECK2_E', 'CHECK3_B',
'CHECK3_E', 'CHECK4_B', 'CHECK4_E', 'Pathogen_type(unique_values)', 'can_cause_sepsis(0:unlikely-
1:probable-9:unknown)', 'pip_resistance(1:sensitive-2:intermediate-3:resistant-9:not_tested)',
'pip_mic(mg/l)', 'target_based_on(0:no_patogen-1:patogen)'
```

- Columns with too many NaN values are dropped:

```
'bicarbonate_v_min', 'bicarbonate_v_max', 'APPLI_E', 'GESDOSIS_N', 'Antimic(0:n-1:y)',
'CHECK1_E(day)',
        'CHECK2_E', 'CHECK3_E', 'CHECK4_E', 'pip_mic(mg/l)'
```

- Columns related to time information were further filtered, together with meta columns:

```
'Sepsis_day', 'Sepsis_hour', 'reason', 'pip_start_hour', 'pip_stop_hour'
'Visite_day_pip_clean_reshape', 'Visite_day_sep_clean_reshape',
          'Start_day_pip_clean_reshape', 'Start_hour_pip_clean_reshape', 'START_D', 'STARTZ_Z',
'STARTGRUND_E',
          'STOPP_D', 'STOPPZ_Z', 'STOPPGRUND_E', 'STOPPGRUND_C',
'Visite_day_ccmc_clean_reshape', 'site',
          'Visite_day_chir_clean_reshape', 'Date_micobio_results', 'Start_day', 'Start_hour',
          'Start_reason(1=>6)', 'STARTGRUND_C', 'Stop_day', 'Stop_time', 'Stop_reason(1=>8)',
'Stop_reason(why_8)',
          'blood_sampling_day', 'blood_sampling_hour', 'why_therapy_stop(1=>8)', 'Stop_pip_day',
'Stop_pip_hour',
          'new_dosage_hour', 'new_dosage_reason(1:TDM-2:side-effect-3:other)', 'KONZENT_D_new',
          'day_visited', 'hospital_admission_date', 'hospital_admission_hour', 'ICU_date', 'ICU_hour',
          'type_of_assigment', 'where_before_ICU'
```

The data cleaning process conducted reduced the number of base features to 100, and number of patients to 253 so far.

Pathogen related information is then encoded. Herein, we created binarized features for each pathogen type, whether they cause sepsis and whether that particular pathogen has pip resistance or not. For that purpose, we filter the pathogen data:

```
['Pathogen_type(unique_values)', 'can_cause_sepsis(0:unlikely-1:probable-9:unknown)',
        'pip_resistance(1:sensitive-2:intermediate-3:resistant-9:not_tested)']
```

where pathogen type can take values of:

```
101, 102, 103, 104, 106, 107, 108, 110, 111, 201, 202, 203, 204, 205, 206, 207, 208, 209, 210, 211, 212,
301, 302, 303, 305, 501, 502, 503, 504, 505, 506, 507, 508, 509, 1051,1052]
```

After data encoding, pathogen information is binarized:

| Pathogen_101(0:n-1:y) | Pathogen_102(0:n-1:y) | Pathogen_103(0:n-1:y) | Pathogen_104(0:n-1:y) |
|---|---|---|---|
| 0 | 0 | 0 | 0 |
| 0 | 0 | 0 | 0 |
| 0 | 0 | 0 | 0 |
| 0 | 0 | 0 | 0 |
| 0 | 0 | 0 | 0 |

with corresponding resistance and sepsis information:

| pip_resistance_508(1:sensitive-2:intermediate-3:resistant-9:not_tested) | pip_resistance_509(1:sensitive-2:intermediate-3:resistant-9:not_tested) | pip_resistance_1051(1:sensitive-2:intermediate-3:resistant-9:not_tested) |
|---|---|---|
| 0 | 0 | 0 |
| 0 | 0 | 0 |
| 0 | 0 | 0 |
| 0 | 0 | 0 |
| 0 | 0 | 0 |

Finally, the remaining dataframe is split as continuous and discrete, based on the number of unique elements in a column. If it is greater than 11, the column is considered to be continuous.

### Pairwise Distance Analysis on Day 1

After the data preparations, without any further dimensionality reduction / feature selection procedure, we checked the similarity between patients in Day 1 in the high dimensional feature space. This is done by computing the pairwise distance between patient i and all remaining patients, followed by an averaging. Figure S2 shows the mean Euclidian distance for each patient. Results revealed that the majority of the patients are at a similar distance to the rest of the patients, whether the patient is part of the control or TDM group. It should be noted that the distances are calculated after scaling, so as not to be biased with respect to large magnitude features.

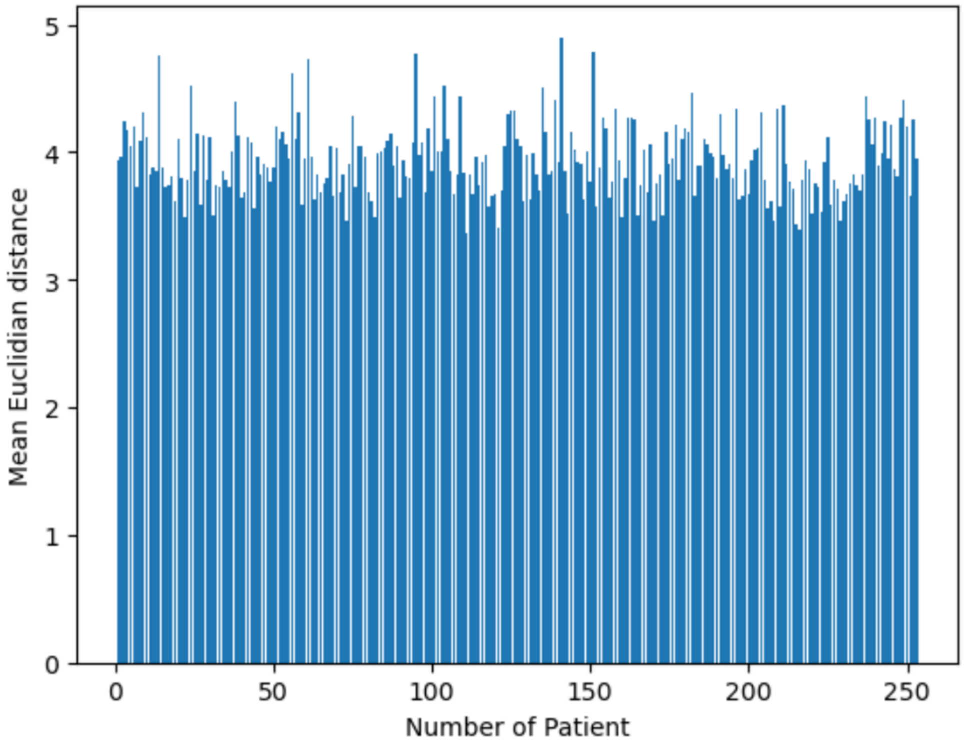


**Figure S2.** Mean pairwise distance in high dimensional feature space between patients for the day of admission.

### Details of Feature Selection via Wrapper

Wrapper methods employ iterative search procedures to select subsets of features for the model. These methods repeatedly provide feature subsets to the model and use the resulting model performance to guide the selection of the next subset for evaluation. The goal is to identify a smaller set of features that outperforms the original set in terms of predictive performance. Wrapper methods offer the advantage of exploring a wider range of feature subsets than simple filters or models with built-in feature selection. However, the main drawback is the excessive computational time required to find the optimal or near optimal subset, particularly if the estimator is a computationally demanding model like neural networks. It should be also noted that wrappers can overfit to the training data, hence requires cross validation-based training procedures.

Wrapper methods can adopt either a greedy or non-greedy approach to feature selection. A greedy search chooses the search path based on the direction that seems the best at the current moment to achieve immediate benefits. While this strategy can be effective, it may reach a locally optimal setting where further improvements become difficult. In contrast, non-greedy search methods such as genetic algorithms (GA) re-evaluate previous feature combinations and have the flexibility to move in a direction that initially appears unfavourable but shows potential benefits in subsequent steps. This allows the non-greedy approach to avoid getting trapped in a local optimum, in which greedy search methods might got caught.

GAs employ a strategy inspired by natural evolution to effectively discover optimal solutions. They generate a set of candidate solutions for the optimization and allow them to reproduce and create new solutions using mating and mutations. Through competition, the most evolutionarily fit solutions, i.e., the optimal ones, have a greater likelihood of surviving and propagating into the next generation (natural selection). This iterative process enables genetic algorithms to gradually improve solutions over time and has demonstrated convergence for a diverse range of problems. For feature selection, the genetic material becomes the indices of feature columns of the original base dataframe, while the length of the genetic material defines the number of selected features. The fitness of a feature subset is calculated via an estimator. Herein, estimator solves a regression task: given the feature set, how accurate the model can predict the labels. In this work, SOFA scores are used as the label and the regression model is chosen to be either a distance-based linear model (multivariate linear regression, Lasso, ElasticNet), or information-based nonlinear models (Random Forests). In other words, we used supervised machine learning methods as the fitness function.

Considering the heterogeneous nature (i.e., continuous, categorical and ordinal features all together) of the patient data, we approached the feature selection with GA in 4 different ways based on (i) the estimator type being used (distance- or information-based) and (ii) whether the data is treated as a whole (199 features passed) or a sub-GA selection is conducted for the continuous, discrete and pathogenic features. For the former, as the distance-based supervised ML models require to work with scaled, continuous features, discrete data is first transformed with CatBoost, and then all features are scaled via a standard scalar. If random forest is used, features are kept as they are; that is, no transformation and no scaling. Secondly, as the dimensionality of the problem is large (199 features), it is considered worthy to investigate a “divide and conquer” approach: a different GA wrapper is used for continuous, discrete and pathogen features, to pick up informative features independently in lower dimensional space. Although lower dimensionality helps to identify patterns in low data limit, this split assumes that continuous, discrete and pathogen feature selections do not have cross correlations. In accordance, we checked SOFA score prediction capabilities based on 4 scenarios:

i. All features are passed to a single GA wrapper as they are (no CatBoost transformation, no scaling) with random forest estimator.
ii. Discrete features are transformed with CatBoost; then all features are scaled with standard scaler. Next, transformed and scaled features are passed to a single GA wrapper with a distance-based linear regression model.
iii. Feature set is split into three; continuous, discrete and discrete pathogen data. These 3 subsets of features are passed to 3 different GA wrapper as they are (no CatBoost transformation, no scaling) with random forest estimator.
iv. Feature set is split into three; continuous, discrete and discrete pathogen data. Discrete and discrete pathogen data are transformed with CatBoost independently; then all three subsets are scaled with standard scaler. Later, transformed and scaled subsets are passed to 3 different GA wrapper with its independent distance-based linear regression model.

For all cases, we first leave 10 patients out completely from the state space database, to leave of some patients for better generalizability. Then, we further conducted a 4:1 train-test split to ensure that feature selection procedure will not leak any information about the evaluation of the state space trajectory analysis. In the next step, we passed the train set of the patient states directly to the GA-based wrapper. Since we did not a priori how many features are enough to represent the patient state, we sweep through number features, starting from 1 to maximum number of features. For instance, in the case of *Feature Set A*, GA Wrapper is called 199 times, for number of features 1,2,3, ..., 198, 199. For each number of features, (e.g., number_of_feature=15), we initialize a population of 160 individuals, with randomly selected 15 features. Herein, the genes of an individual contain the indices of 15 randomly picked features. Then, for each individual, a different estimator is being trained with a cross validation scheme (cv=5). The negative mean absolute error of that individual's cv scores is saved as the fitness. Then, the same is applied to all 160 individuals. This is followed by a parent selection, mating and mutation, which enables to update the gene pool. The whole GA process is applied for 100 generations, yielding the best feature sub-set (e.g., best 15 sub-features among 199). Lastly, the trained estimator with the best feature subset is used to generate repeated k-fold cv scores (RepeatedKFold(n_splits=10, n_repeats=3). Scores are saved. The process continues with the next number of features (e.g., number of features = 15+1=16) until the maximum number of features is reached.

Figure S3 shows the repeated k-fold scores (negative MAE) with increasing number of features for scenario A. The predictive accuracy of the feature subset yielded such plateaus in all feature selection scenarios A-D, indicating that adding more features do not contribute to the model accuracy after a threshold, due to the limitations dictated by the sparsity of the dataspace. What differed between scenarios A-D was the negative MAEs, and the number of features it yields for its maximum accuracy. Overall, scenario A yielded the highest accuracy at lower number of features.

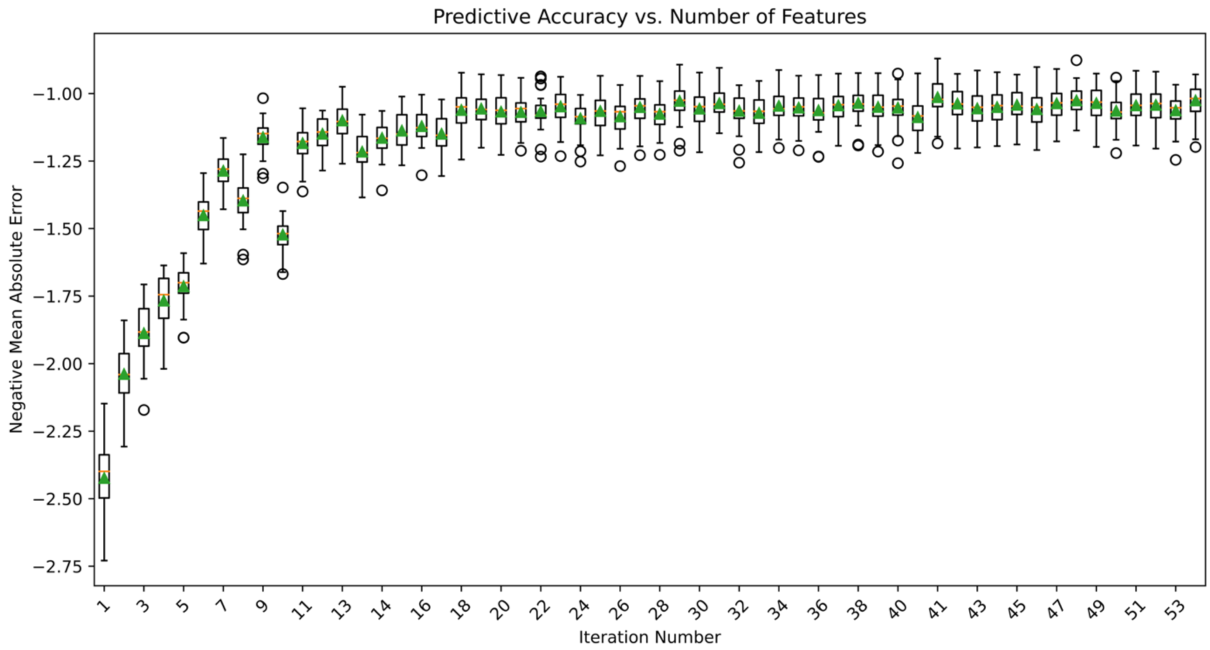


**Figure S3.** Predictive accuracy of the estimator with increasing number of features. For Scenario A, y axis denotes the -MAE of predicting the patient's SOFA score given the feature subset in repeated k-fold analysis.

After determining the number features needed to accurately represent the patient state as 23 from **Figure S3** for Scenario A, we run the GA wrapper to pick the best 23 feature subsets 100 times. Variations in the predictive accuracy for each run is depicted in **Figure S4**. This analysis further enable us to investigate the frequency of features being selected during the stochastic evolutionary process.

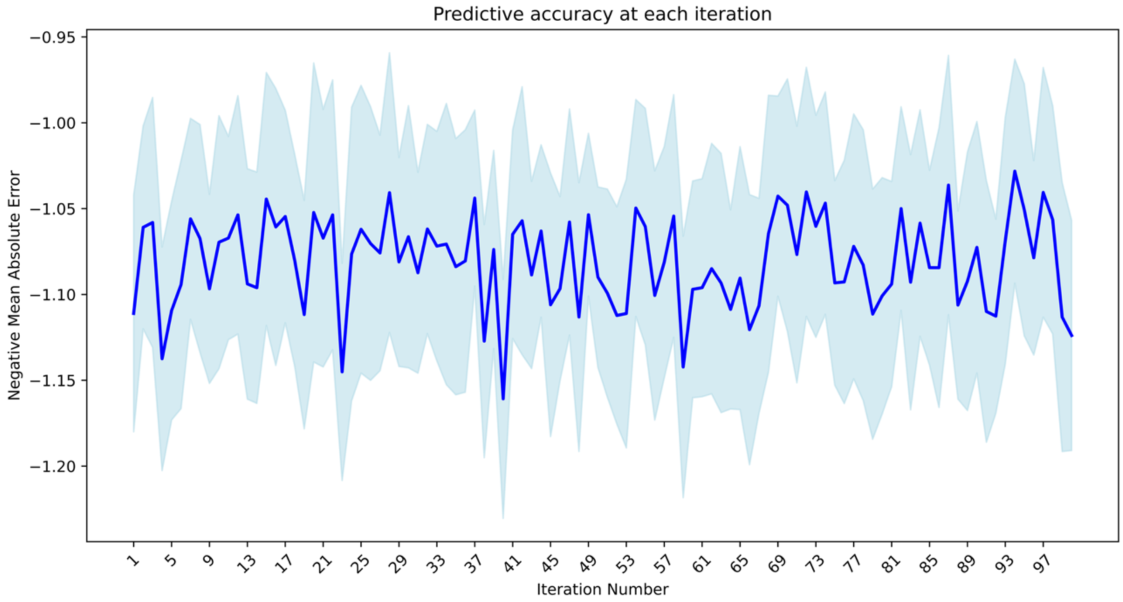


**Figure S4.** Variations in repeated k-fold analysis for feature selection with GA (number_of_feature = 23). Each point shows the -MAE of the last generation of each 100 run.

Another interesting observation worth mentioning is the impact of estimators with implicit feature selection properties (i.e., Lasso, ElasticNet) once they are used within the wrapper. Herein, we observed a systematic decrease in predictive accuracy if linear regression models are used with regularization within the wrapper. **Figure S4** depicts the trend for scenario C, continuous data subset, where the number of features is taken at the left limit of its own plateau curve.

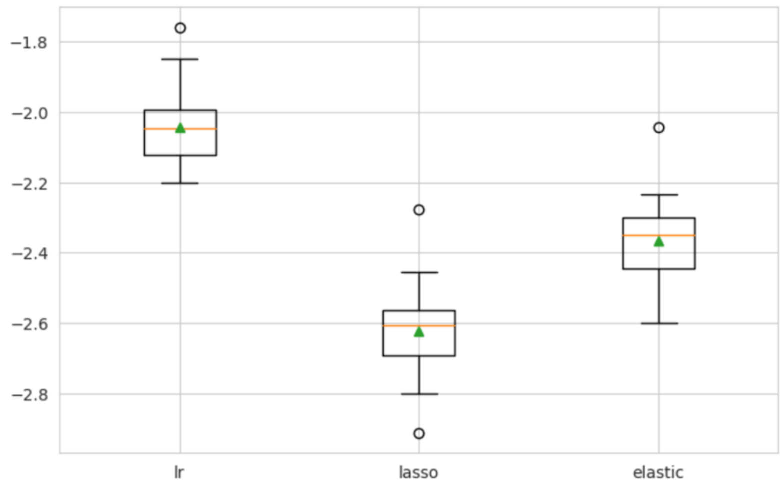


**Figure S5.** Impact of using implicit feature selector as an estimator in GA wrapper. y axis denotes the -MAE in SOFA score predictions. lr: linear regression.

### SOFA Score Analysis

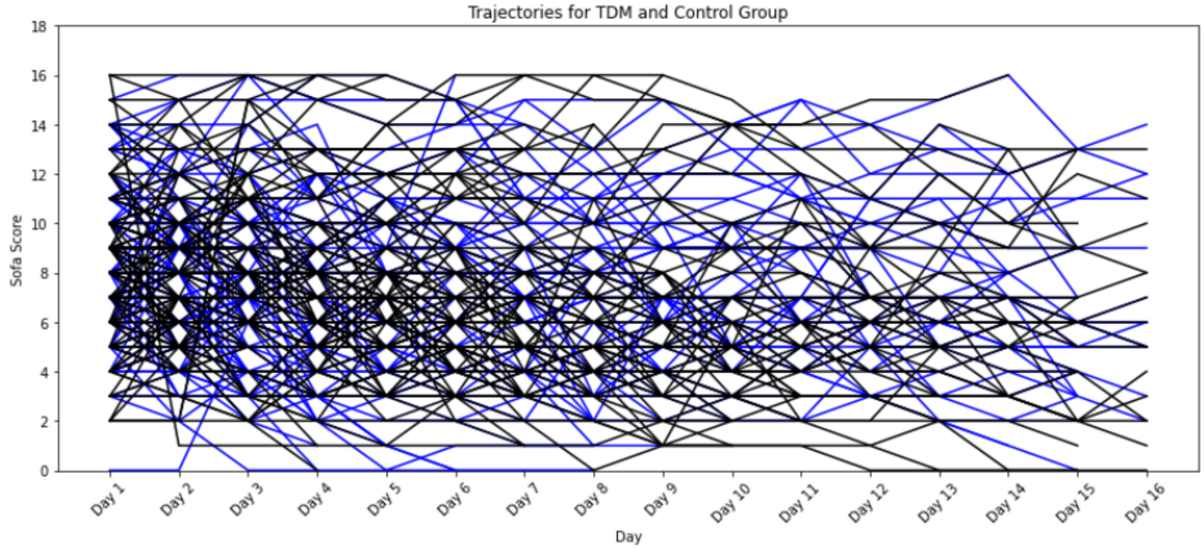


**Figure S6.** Fluctuations in patient SOFA score trajectories. Patient health state jumps significantly between SOFA scores on a daily basis. Each curve represents a patient. Blue: TDM, Black: control.
